# Activity Space Maps: a novel human mobility data set for quantifying time spent at risk

**DOI:** 10.1101/2021.08.01.21261352

**Authors:** Daniel T. Citron, Shankar Iyer, Robert C. Reiner, David L. Smith

## Abstract

Activity Space Maps are a novel global-scale movement and mobility data set which describes how people distribute their time through geographic space. The maps are intended for use by researchers for the purposes of epidemiological modeling. Activity Space Maps are designed to complement existing digitally-collected mobility data sets by quantifying the amount of time that people spend in different locations. This information is important for estimating the duration of contact with the environment and the potential risk of exposure to disease. More concretely, the type of information contained in Activity Space Maps will make it easier to model the spatial transmission patterns of vector-borne diseases like malaria and Dengue fever. We will discuss the motivation for designing Activity Space Maps, how the maps are generated from mobile phone user app location history data, and discuss an example use case demonstrating how such data may be used together with spatial epidemiological data to advance our understanding of spatial disease patterns and the relationship between travel behaviors and infection risk.

## 1 INTRODUCTION

Data sets which describe human movement patterns are important for understanding epidemiologically relevant interactions and contact patterns. Most disease-causing pathogens are not able to move great distances on their own. Instead, the pathogens can be transported between places by human and animal hosts. Infected people may travel from their home territory to another location and infect others there, causing a new outbreak; or, travelers may contract an illness while away and unintentionally spread it to their households and neighbors after returning home. Knowing where it is that people go and how they move around in geographical space is key for mapping out spatial patterns of contact and exposure [16, 17, 27].

Until recently, it was very difficult to obtain accurate data on human movement patterns at scale. Medical surveys of patients are subject to selection bias (they only survey people who end up at a healthcare facility) and patient recall bias (may not accurately report their travel history) [13]. Distributing surveys to the general population is expensive to accomplish at scale [4] and similarly affected by recall biases. More recently, it has become possible for researchers to distribute GPS location history trackers to people as a way of recording their movements, but these studies are hard to sustain at a large scale [19]. With the widespread adoption of mobile phones, digitally-collected mobile-phone user location history data offers an unprecedented opportunity to quantify patterns of movement behavior at a broad scale [1, 11, 14, 25]. Effectively, this becomes an observational study similar to the GPS location history tracker studies, but without the need to distribute and maintain devices.

The formatting of digitally-collected location history data matters for its applicability for research. Many early movement data sets have been aggregated and formatted as origin-destination matrices, which count the number of users who travel from one location to another during an observation period [12]. This formatting is useful both for parsimony as well as for disguising individual user trajectories for the sake of preserving privacy. While valuable for understanding population flows, data formatted as origin-destination matrices do not contain information on how much time a traveler spends in any location. For example, the matrix might count one user passing from location A to location B on a particular day, but not know whether the user spent the entire day in location B or whether they immediately traveled from location B to location C. For epidemiological purposes, the duration of time that a user spends in any given location is key for understanding epidemiologically relevant contact and exposure patterns [2, 6, 23]. After all, a person who spends only a day in a setting where they are at risk of contracting malaria is less likely to become sick than if they spend two weeks in that setting.

## 2 DATASET DESCRIPTION

Activity Space Maps are a novel data set developed through Facebook’s Data for Good initiative. They are built using de-identified Facebook mobile-app user location history data of users who consented to share their precise location and opted into Facebook’s location history collection. The maps aggregate movement trajectories across space and time such that they represent a probability distribution, describing the average amount of time that a user from one location spends visiting other locations. That is to say, what is the probability that an average individual spends in one location, given that their home residence is found in another location. The maps are formatted similarly to the space utilization functions that are found in ecology which are constructed using GPS trackers and aggregated as a way of summarizing how it is that animals move around and use space [9].

Activity Space Maps are generated by collecting and aggregating individual users’ location histories. All users included in the pipeline have consented to collection of their precise location and have opted into Facebook’s location history collection. The raw source data are location history traces, initially recorded as a series of tuples of the form user, time, location. Initially, locations are binned using Microsoft’s Level 12 Bing tile maps, a quadkey map of square regions approximately 9.8 kilometers to a side at the equator [18]. Each user’s trajectory is binned into a series of 24-hour periods. Each 24-hour period is denoted by a date stamp and contains one nighttime period, spanning from 8 p.m. on the previous date to 6 a.m. on the given date, and one daytime period, spanning from 8 a.m. to 6 p.m. on the given date. For each date, modal locations for both the daytime and nighttime periods are extracted. At this stage, all users who do not consistently use location history services and all users who appear in the data set fewer than 7 out of the past 28 nights are excluded. The next step is to aggregate the total number of person-periods (i.e., person-days and person-nights) spent by each individual in each location across the last 28 days. This protects users’ privacy by removing links between specific locations and specific visit times and instead summarizes each user’s space utilization over the previous four weeks. Each user’s home location is estimated using the most frequently visited nighttime modal location during the 28-day aggregation period. The assumption here is that, while there are some instances of people spending their nights away from their home residences, the vast majority of people are most likely to sleep at home on most nights.

Lastly, Activity Space Maps are generated by aggregating over all users in each home location, producing a probability distribution showing how the average individual who lives in a particular location distributes their time across geographical space. The resulting dataset quantifies, for each home-visit tile pair, the relative fraction of person-days and person-nights spent in the visit tile location by users who live in the home tile location. In this way, Activity Space Maps quantify typical movement behavior and patterns of users who live in the same location. Figure 1 shows two example plots of Activity Space Maps data collected for a home tile in Rome, Italy. Both the daytime and nighttime spent distributions are shown. The temporal aggregation means that the dataset reflects daily, weekly, or monthly movement patterns such as commuting or short-term trips. The data set is not intended to capture or reflect longer-term patterns such as seasonal migration or permanent relocation.

**Figure 1:**
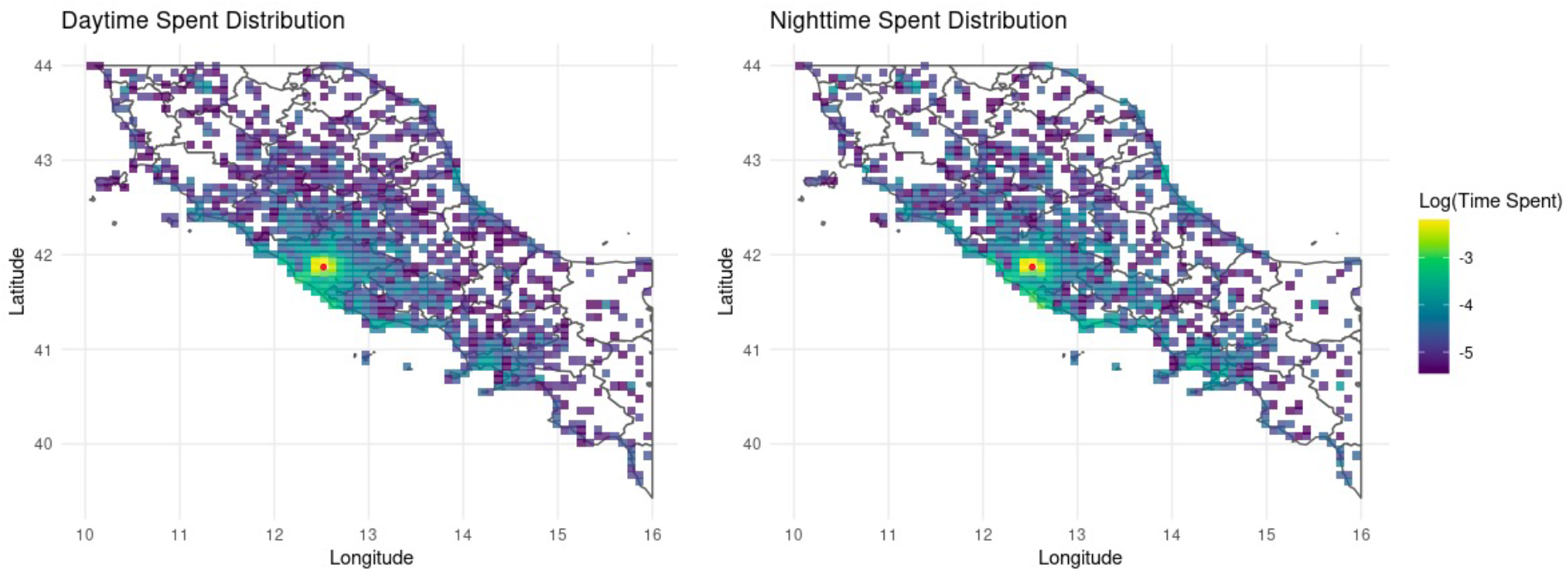
Activity Space Maps example: The data plotted in the two maps were collected for a home tile in Rome, Italy during the 28-day period ending on June 15th, 2021. The map on the left shows the daytime spent distribution and the map on the right shows the nighttime spent distribution. The red dot indicates the home location. The other colors denote the logarithm of the relative probability of spending a day or night in each other location.

Preserving individual user privacy is a key feature of Activity Space Maps and all Facebook Data for Good products. No individual trajectories or records which pinpoint any individual’s visit to a location at a particular time are exposed through this data set. All locations which don’t meet a population density threshold are excluded from the data set for the sake of concealing any individual trajectories. Following guidance from humanitarian organizations who have partnered with Facebook Data for Good, cross-border movements are excluded from the dataset for the sake of protecting at-risk populations who move in times of crisis.

Activity Space Maps collect data in 151 countries and territories worldwide. In terms of the number of users sampled, Activity Space Maps has the most coverage for countries in Europe, North America, South America, South Asia, and Southeast Asia, but less coverage in other parts of Asia, Africa, and Oceania. Activity Space Maps are recorded on an ongoing weekly basis, beginning in mid-April of 2021. Facebook Data for Good plans to release Activity Space Maps to vetted Data for Good external partners in the second half of 2021.

We must highlight a few properties of how Activity Space Maps are constructed which have the potential to skew or bias the movement patterns which they represent. Primarily, it is likely that the sampled population does not represent an unbiased sample of the overall population [25, 26]. The data only reflect those who have opted into location history services through the Facebook mobile app - it is plausible to think that such individuals may have different movement patterns than those who do not access these features. Furthermore, the data set is only able to represent the movements of people who own mobile phones while they are traveling in areas with access to phone service. For these reasons, we caution users that the data sets are likely to over-represent the movements of people who live in higher-income, urban areas and under-represent the movements of people who live in lower-income, rural areas. Secondly, there is also the potential problem of under-sampling in low population density areas, combined with the privacy-related exclusion of low density areas. For this reason, Activity Space Maps currently are able to reveal the connectivity patterns originating in high-density, urban areas but are less likely to portray connectivity patterns originating in low-density, rural areas. Finding methods to account for and correct potential sources of bias in the data is an ongoing line of research, both within the Facebook Data for Good team as well as within the broader research community [10, 28].

## 3 APPLICATIONS

There are a variety of potential epidemiological applications for Activity Space Maps. Generally speaking, Activity Space Maps have a potential for improving understanding of epidemiological patterns where movement and travel expose people to hazards in the environment. To give one example, we outline research which combines Activity Space Maps with malaria prevalence maps from the Malaria Atlas Project (MAP) [5, 24] using a mechanistic transmission model to show how travel drives the incidence of malaria among people living in urban areas in sub-Saharan Africa. This line of research draws on earlier analysis performed for Bioko Island in Equatorial Guinea [3, 7, 8], which estimated the fraction of malaria prevalence which may be attributable to exposure to infectious mosquitoes while traveling.

Malaria is a vector-borne parasitic disease which primarily affects people living in tropical climates in sub-Saharan Africa, Asia, and Central and Southern America. The WHO estimates that over 220 million malaria cases and 400,000 deaths due to malaria occurred in 2019 alone, making it one of the most dangerous causes of death and disease [15]. Malaria cases primarily affect children and people living in rural settings. Malaria is spread by *Anopheles* mosquitoes, who pass parasites to human hosts when they blood feed. Because malaria is transmitted by an insect vector, the risk experienced by people depends strongly on whether or not those people spend time in areas where infectious vector mosquitoes live: one will be at risk of becoming infected if one spends time in areas where the local environment supports a viable vector population. It is for this reason that knowing where people spend their time is so important for understanding who is at risk and the patterns of transmission.

The Malaria Atlas Project (MAP) produces raster maps of malaria prevalence, or parasite rate (PR) [5, 24], showing the proportion of infected people in a given location. The maps reveal the geographical heterogeneity of malaria prevalence, which reflects the geographical heterogeneity in the local disease-producing ecology as experienced by human hosts. The PR maps show where it is that infected people are located, but the maps do not show where it is that the people became infected. In many cases, most likely people are becoming infected in their own homes. However, some of those cases are likely due to exposure that occurred during travel. Malaria vector mosquitoes typically do not thrive in urban areas. It is likely that cases of malaria reported in urban areas are attributable to travel.

Bioko Island represents one setting where this is likely to be the case. Since 2004, the Bioko Island Malaria Elimination Program has implemented an extensive program of interventions against malaria [4]. Despite this, malaria persists among island residents, most of whom live in the urbanized capital city of Malabo. Using travel data collected through surveys in 2015-2018, it was estimated that much of the malaria occurring in Malabo could be attributable to travel to mainland Equatorial Guinea, where there is a high risk of malaria [3, 7, 8]. This is an example of how travel data may be used to discover source-sink disease dynamics, where the high incidence of disease in one location drives the occurrence of disease in another location because of people traveling from one location to another [16, 17].

Activity Space Maps may be used to perform similar analysis, this time using digitally-collected data to infer travel patterns. The leftmost plot in Figure 2 shows the Activity Space Map for individuals living in the highlighted location in Lagos, Nigeria (marked with a red dot). The distribution plotted shows that most time is spent in the area of Lagos, but that some trips are taken to Abuja and the Niger river delta region. The center plot in Figure 2 shows the Malaria Atlas Project’s estimated prevalence levels of malaria (PR) in 2015 among children aged 2-10, plotted only in the same areas for which Activity Space Maps also provides data. The maps show how dense urban areas like Lagos and Abuja tend to have lower prevalence, but that there still are some infected people living in these areas.

**Figure 2:**
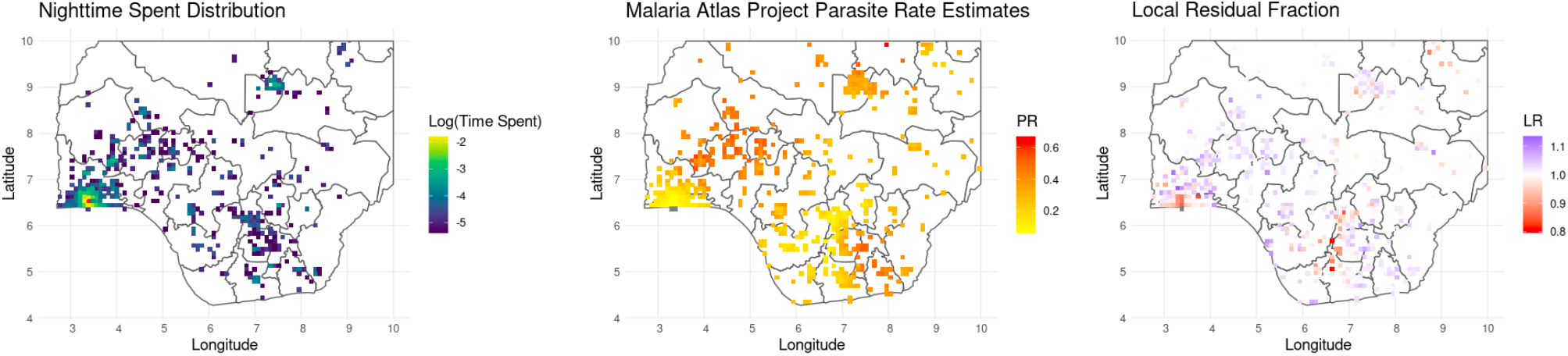
Example analysis: Left plot illustrates the distribution of nighttime periods spent by residents in part of Lagos, Nigeria (red dot indicates home location), with the color scale showing the logarithm of the relative probability of spending a night in each visit location. Center plot shows Malaria Atlas Project *Plasmodium falciparum* Parasite Rate (PR) estimates for children aged 2-10 in Nigeria in 2015. Right plot shows the local residual fraction of prevalence which is attributable to local transmission. This last plot is the result of analysis which uses Activity Space Maps to estimate the influence which travel has on local transmission. The areas marked in red are locations where malaria exposure occurs when people travel to other areas, and the areas marked in purple are locations where malaria exposure mostly occurs locally.

By combining together the epidemiological prevalence data and the movement data, it is possible to infer where it is likely that people are becoming exposed and infected. The next step of the analysis utilizes on a mechanistic model of malaria transmission [3, 16, 17, 20]. The model relates the prevalence to the force of infection (FOI) experienced by individuals in different locations. The FOI is a way of quantifying risk to individuals, an infection rate which reflects exposure to infectious vectors in the local environment. Within the model, the average individual’s overall FOI is expressed as a weighted average of the amount of time spent in each location visited, multiplied by the FOI in each location. Beginning with the prevalence map, inverting the model results in an estimate of local FOI in each location on the map.

Knowing the true FOI in each location makes it possible to estimate the exposure rate experienced by each individual in each place they travel to. Activity Space Maps describe how an average individual distributes their time through geographical space, making it possible to derive estimates of where it is that people are exposed to malaria. The rightmost plot in Figure 2 plots Local Residual Fraction, or the fraction of prevalence that is attributable to local transmission at one’s home location [3]. The areas marked in red, such as in Lagos, are areas where exposure that occurs while traveling has a significant impact on local malaria prevalence. That is to say, people with malaria may have been exposed elsewhere. The areas marked in purple, such as in the region to the north of Lagos, are areas where local transmission is strong and travel does not have much influence on overall prevalence. This simple preliminary analysis can provide some insights into where risk of infection occurs, and can assist with strategic planning for intervention programs. The results suggest that there are some areas where interventions to decrease local exposure should be prioritized, but also that there are other areas where additional protections for travelers could be helpful.

Replicating the analysis of travel and malaria seen in the Bioko Island study in a large country using traditional survey data would be very costly, but Activity Space Maps has made it possible to perform a spatial analysis of transmission patterns for Nigeria using digitally-collected data. This analysis represents one example of how the movement maps may be applied to improve understanding of spatial patterns of disease transmission. Generally speaking, Activity Space Maps are designed to understand how movement patterns bring people into contact with hazards in the environment. Activity Space Maps may be able to support research on other vector-borne diseases, such as Dengue fever [21, 22]. For water-borne diseases such as cholera, exposure occurs through a different mechanism, but Activity Space Maps may provide insights into identifying which populations spend time in places where they are at risk. Lastly, there is potential for applications related to directly transmitted pathogens such as SARS-CoV-2 [1, 14], where Activity Space Maps may support existing movement data sources to understand which populations travel to high-transmission locations.

## Data Availability

Facebook Data for Good plans to release Activity Space Maps to vetted Data for Good external partners in the second half of 2021. Details related to the mathematical modeling exercise will be provided upon request by email.

